# Integrative Omics Reveal Novel Protein Targets For Chronic Obstructive Pulmonary Disease Biomarker Discovery

**DOI:** 10.1101/2021.01.11.21249617

**Authors:** Ana I Hernandez Cordero, Stephen Milne, Chen Xi Yang, Xuan Li, Henry Shi, Don D. Sin, Ma’en Obeidat

## Abstract

**Background:** Large genome-wide association studies (GWAS) and other genetic studies have revealed genetic loci that are associated with chronic obstructive pulmonary disease (COPD). However, the proteins responsible for COPD pathogenesis remain elusive. We used integrative-omics by combining genetics of lung function and COPD with genetics of proteome to identify proteins underlying lung function variation and COPD risk.

**Methods:** We used summary statistics from the GWAS of human plasma proteome from the INTERVAL cohort (n=3,301) and integrated these data with lung function GWAS results from the UK Biobank cohorts (n=400,102) and COPD GWAS results from the ICGC cohort (35,735 cases and 222,076 controls). We performed in parallel: a proteome-wide Bayesian colocalization, and a proteome-wide Mendelian Randomization (MR) analyses. Next, we selected proteins that colocalized with lung function and/or COPD risk and explored their causal association with lung function and/or COPD using MR analysis (*P*<*0*.*05*).

**Results:** We found 537, 607, and 250 proteins that colocalized with force expiratory volume in one second (FEV_1_), FEV_1_/forced vital capacity (FVC), or COPD risk, respectively. Of these, 1,051 were unique proteins. The sRAGE protein demonstrated the strongest colocalization with FEV_1_/FVC and COPD risk, while QSOX2, FAM3D and F177A proteins had the strongest associations with FEV_1_. Of these, 37 proteins that colocalized with lung function and/or COPD, also had a significant causal association. These included proteins such as PDE4D, QSOX2 and RGAP1, amongst others.

**Conclusion:** Integrative-omics reveals new proteins related to lung function. These proteins may play important roles in the pathogenesis of COPD.

## Introduction

Chronic obstructive pulmonary disease (COPD) is a persistent respiratory condition that is characterized by irreversible and progressive lung function impairment, and is responsible for over three million deaths worldwide each year (Roth et al. 2018). Large genome-wide association studies (GWAS) have revealed hundreds of genetic loci that are associated with COPD (Cho et al. 2014; Hobbs et al. 2017; Sakornsakolpat et al. 2019) and lung function more generally (Shrine et al. 2019). However, the molecular mechanisms relating these genetic associations to lung function and COPD pathogenesis remain elusive, hindering the ability to translate these findings into new therapeutic targets or biomarkers of disease.

Integrative-omic methodologies may provide insights into the biological relationships between genetic variants and complex traits such as lung function and COPD (Giambartolomei et al. 2014; Gusev et al. 2016). These methods aim to establish a link between gene expression or protein levels and the trait by leveraging their respective associations with common genetic variants, which can be determined from independent cohorts. For example, the integration of COPD GWAS with transcriptomic datasets suggests that the effects of many COPD risk loci are mediated through regulation of gene expression in lung tissue (Obeidat et al. 2015; Lamontagne et al. 2018).

Determining causal associations between molecular and complex traits are critical for understanding disease pathogenesis, and for translating these molecules into biomarkers or therapeutic targets. One method that has recently gained momentum in assessing causality of a complex trait-molecular phenotype relationship is Mendelian Randomization (MR). The MR framework exploits the random allocation of alleles during meiosis and relates their effects on a putative risk factor, which can be a quantified by measuring biomolecules such as a blood protein (Smith and Ebrahim 2003; Voight et al. 2012). This in turn can be related to a trait. MR analysis measures the ‘lifetime exposure’ to this risk factor in a way that is relatively resistant to confounding from environmental influences or reverse causation. This enables an unbiased assessment of causality. MR analysis has established causal associations between a number of candidate blood proteins and COPD (Milne et al. 2020). However, the application of MR analysis at a genome-wide discovery level and by coupling it with integrative omics methods will likely yield many additional novel protein associations with COPD.

In this study, we combined two approaches; MR analysis of plasma proteins and COPD risk and genome-wide Bayesian colocalization (COLOC). We used both of these approaches to integrate a large human plasma proteome dataset (Sun et al. 2018) with GWASs for lung function in a general population (Shrine et al. 2019) and for COPD risk in a large case-control dataset (Sakornsakolpat et al. 2019), to nominate promising protein targets for further exploration in mechanistic and biomarker studies.

## Results

### Study design

The overall study design is shown in **Figure 1**. We aimed to identify plasma proteins causally associated with the following phenotypic traits: forced expiratory volume in one sec (FEV_1_); FEV_1_ to forced vital capacity (FEV_1_/FVC) ratio; and the presence of COPD (henceforth referred to as ‘COPD risk’). These associations were determined by integrating a human plasma proteome GWAS (Sun et al. 2018) with the largest existing GWAS for lung function (Shrine et al. 2019) and COPD (Sakornsakolpat et al. 2019). Each of these datasets (described in detail in the Methods) summarizes the association of millions of genetic variants (single nucleotide polymorphisms [SNPs]) with their respective traits. We performed the study in two stages. Stage 1 was an unbiased discovery of proteins associated with the phenotypic traits, by performing two integrative omics methods in parallel: a genome-wide COLOC analysis of each associated genetic locus, and a genome-wide MR analysis of all measured proteins. Stage 2 was a step-wise analysis, wherein we used only proteins that were significantly colocalized with one or more of the phenotypic parameters in subsequent, apply a hypothesis-driven MR threshold. From these results, we generated a list of plasma proteins that showed a causal association with COPD risk and/or lung function.

**Figure 1.**
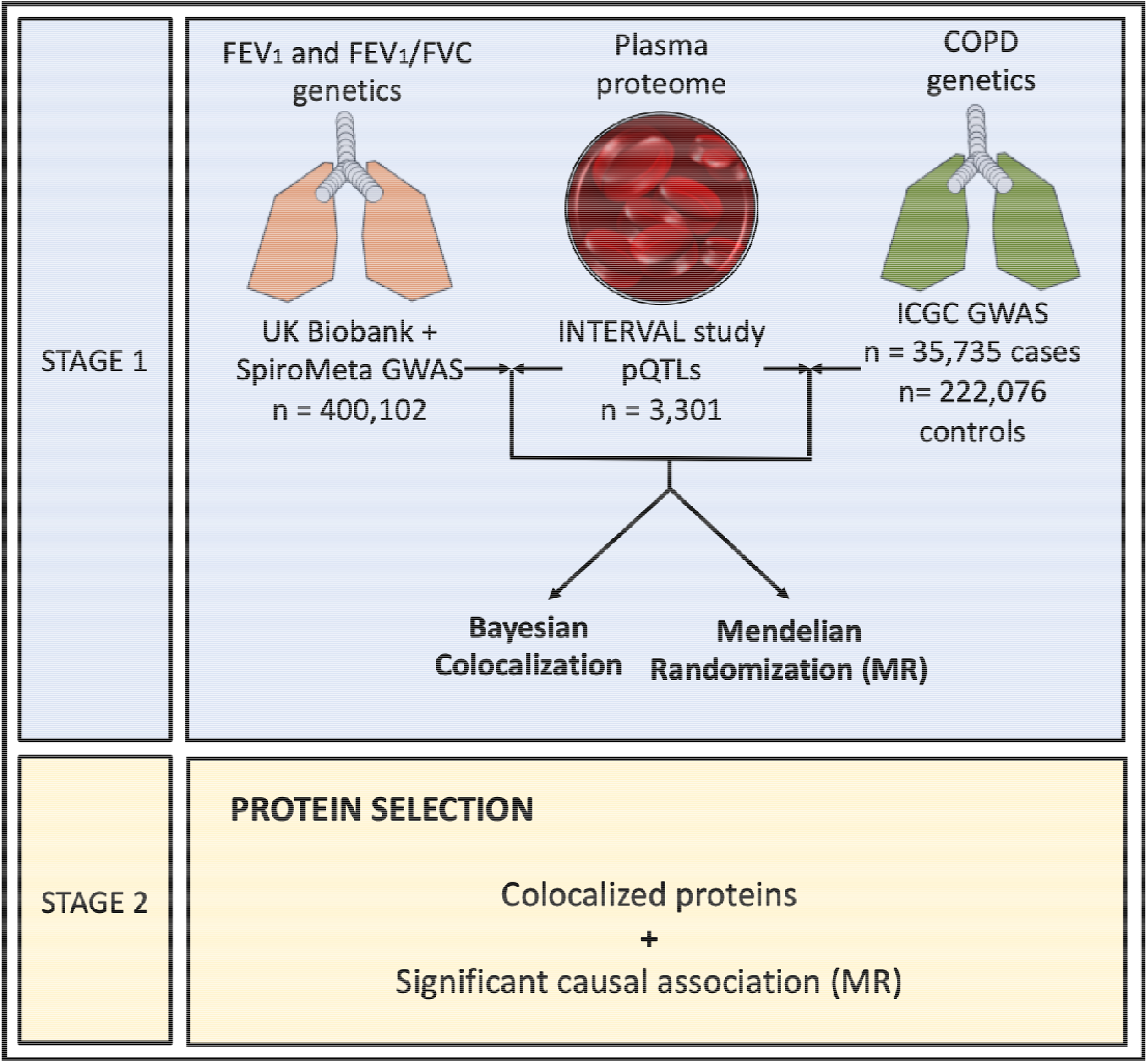
Study design. The diagram shows the workflow used to identify causal factors for lung function and COPD risk. We selected as top plasma proteins those that showed significant colocalization at *PP*_*H4*_ > 0.80 and causality at *FDR* < 0.10 or *P*_*MR*_ < 0.05 with lung function and/or COPD risk.

### Stage 1: a genome-wide discovery of plasma proteins associated with lung function and COPD risk

In Stage 1, we used two integrative omics methods. We first performed a COLOC analysis across the 2,995 proteins, which were measured in the plasma proteome dataset. For this analysis, we included genetic loci associated with each of the clinical parameters (lung function traits and COPD) at a *P*_*GWAS*_ < 5×10^−07^ and set the significance of colocalization at *PP*_*H4*_ > *0*.*80*.

In total, 1,048 unique proteins were colocalized with at least one of the COPD phenotypes. Of these, 447 protein colocalized at *PP*_*H4*_ > *0*.*90*. For the lung function traits, 537 proteins colocalized with FEV_1_; proteins with the highest *PP*_*H4*_ were sulfhydryl oxidase 2 (QSOX2) (*PP*_*H4*_ *= 0*.*99*), protein FAM3D (FAM3D) (*PP*_*H4*_ *= 0*.*99*) and FAM177A1 (F177A) (*PP*_*H4*_ *= 0*.*99*) (**Figure 2**). Likewise, 607 proteins colocalized with FEV_1_/FVC; those with the highest *PP*_*H4*_ included the advanced glycosylation end product-specific receptor (sRAGE) (*PP*_*H4*_*= 0*.*99*), stromelysin-2 (MMP-10) (*PP*_*H4*_ *= 0*.*99*) and collagen alpha-3(VI) (*PP*_*H4*_ *= 0*.*99*) (**Figure 2**). We found that 200 unique proteins overlapped with both traits (**Supplemental Table 1**). For COPD risk, there were 250 colocalized proteins. Based on *PP*_*H4*_, the top three colocalized proteins were sRAGE (*PP*_*H4*_ *= 0*.*99*), C-C motif chemokine 14 (HCC-1) (*PP*_*H4*_ *= 0*.*99*), and AT-rich interactive domain-containing protein 3A (ARI3A) (*PP*_*H4*_*=0*.*99*) (**Figure 2**). Approximately half of proteins that were colocalized with COPD risk were also colocalized with FEV_1_ (94/250) or FEV_1_/FVC (126/250) (**Supplemental Table 1**).

**Table 1.**
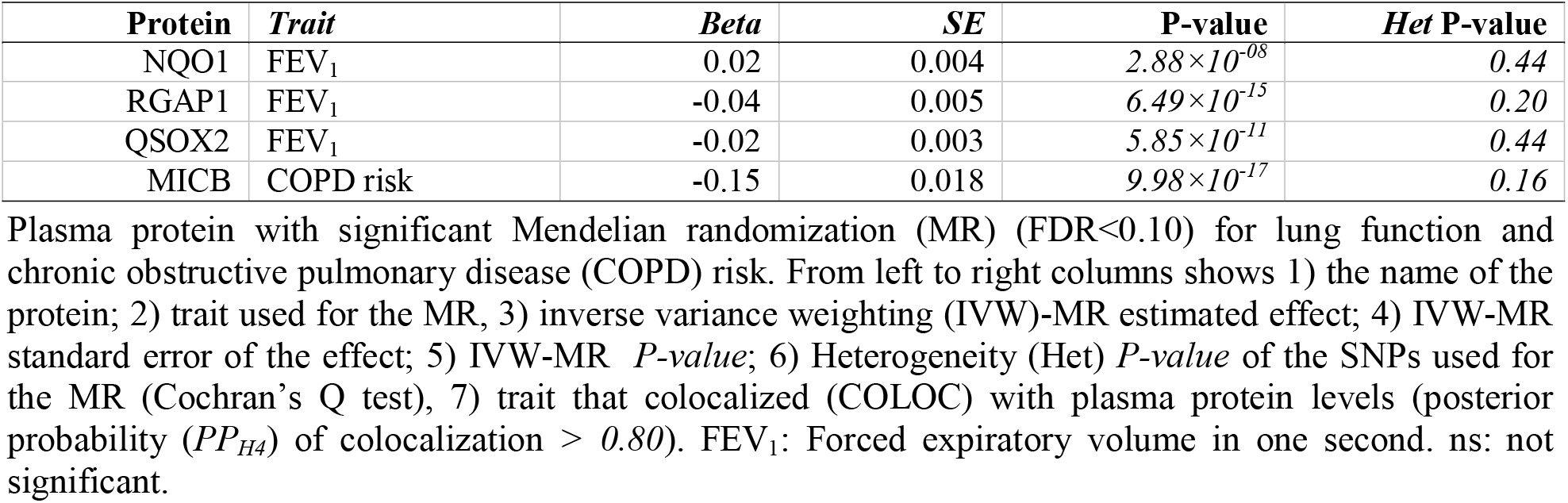
**Causal association of plasma proteins with lung function and/or COPD risk from hypothesis-free MR analysis**

| Protein | Trait | Beta | SE | P-value | Het P-value |
| --- | --- | --- | --- | --- | --- |
| NQO1 | FEV<sub>1</sub> | 0.02 | 0.004 | $2.88 \times 10^{-08}$ | 0.44 |
| RGAP1 | FEV<sub>1</sub> | -0.04 | 0.005 | $6.49 \times 10^{-15}$ | 0.20 |
| QSOX2 | FEV<sub>1</sub> | -0.02 | 0.003 | $5.85 \times 10^{-11}$ | 0.44 |
| MICB | COPD risk | -0.15 | 0.018 | $9.98 \times 10^{-17}$ | 0.16 |

Plasma protein with significant Mendelian randomization (MR) ( $FDR < 0.10$ ) for lung function and chronic obstructive pulmonary disease (COPD) risk. From left to right columns shows 1) the name of the protein; 2) trait used for the MR, 3) inverse variance weighting (IVW)-MR estimated effect; 4) IVW-MR standard error of the effect; 5) IVW-MR $P$ -value; 6) Heterogeneity (Het) $P$ -value of the SNPs used for the MR (Cochran's Q test), 7) trait that colocalized (COLOC) with plasma protein levels (posterior probability ( $PP_{H4}$ ) of colocalization $> 0.80$ ). FEV<sub>1</sub>: Forced expiratory volume in one second. ns: not significant.

**Figure 2.**
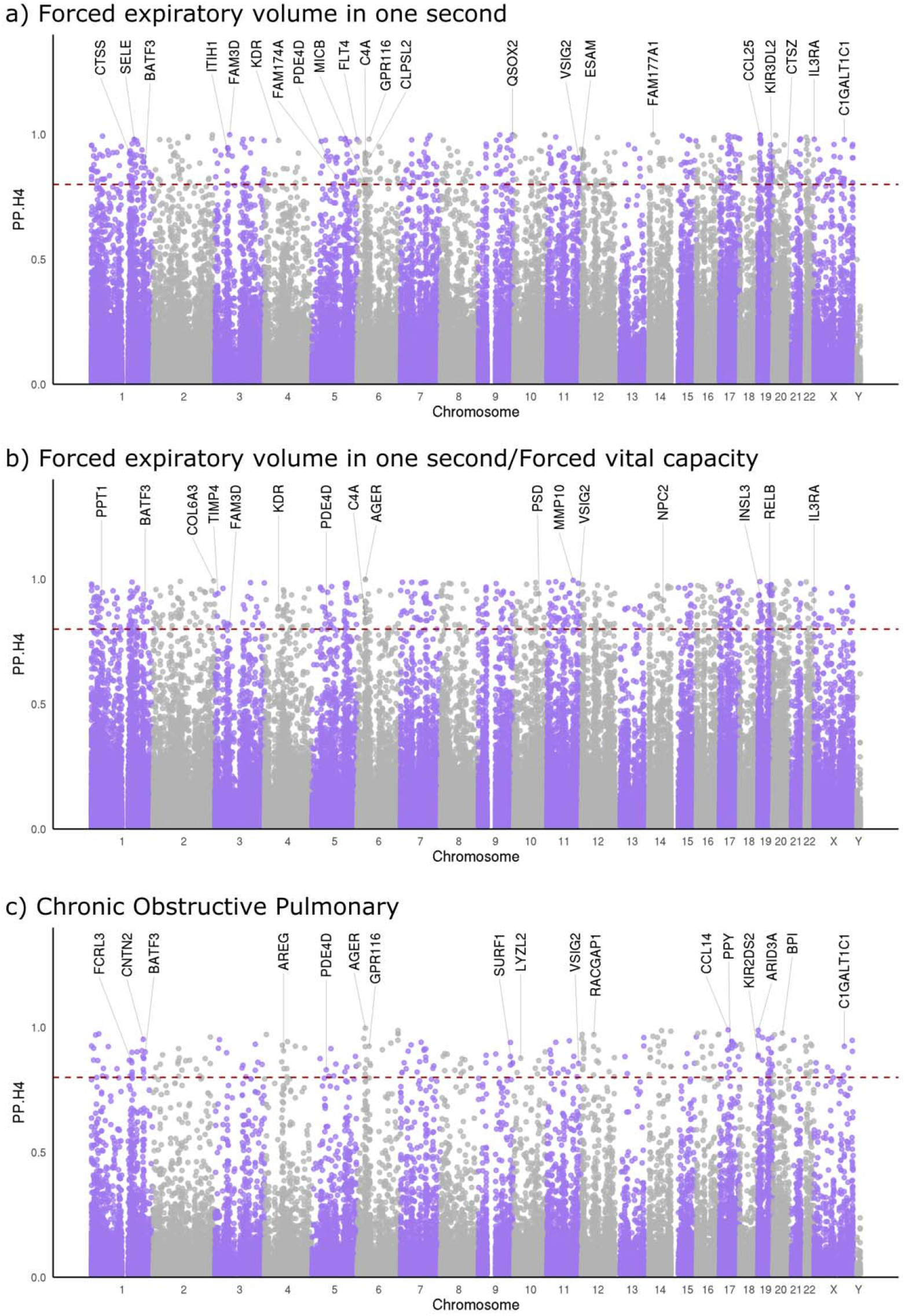
Bayesian colocalization (COLOC) analyses of the plasma proteome related to lung function (a and b) traits and chronic obstructive pulmonary disease (COPD) risk (c). The horizontal axis in each plot represents the chromosomal position of the plasma protein coding genes and the vertical axis shows the posterior probability of the two phenotypes (protein level and clinical trait) sharing common genetic variant (PP_*H4*_). Significant colocalization is defined as *PP*_*H4*_ >0.80 (red dashed line). Each purple or grey dot represents a plasma protein. Labelled colocalized proteins are those with significant causal associations with the phenotypic trait at *P*<*0*.*05*. Note: Plasma proteins are labelled based on their protein coding gene names.

We next performed MR analyses for each of the 2,995 proteins present in the plasma proteome dataset (Sun et al. 2018). MR analysis uses genetic variants as instrumental variables to relate their per-allele effects on a risk factor to their per-allele effects on a phenotypic trait. MR therefore estimates the ‘causal’ effect of the risk factor on the phenotypic trait. For these analyses, we used a two-sample multivariable inverse variance weighted MR model (IVW-MR), and tested various MR assumptions (see Methods). Since this was a hypothesis-free analysis, we set a false discovery rate (FDR) of <0.1 to control for multiple comparisons.

For the lung function traits, three proteins – QSOX2, Rac GTPase-activating protein 1 (RGAP1) and NAD(P)H quinone dehydrogenase 1 (NQO1) – showed causal associations with FEV_1_ at FDR < 0.1 (**Table 1**). Testing of the MR assumptions showed no significant heterogeneity based on a Cochran’s Q test (*P* > *0*.*05*), or pleiotropy (Egger-MR intercept *P* > *0*.*05*) for either protein. Based on the direction of the MR estimate, we inferred that increased plasma levels of QSOX2 and RGAP1 were associated with decreased FEV_1_ (**Table 1 and Figure 3**), while increased plasma level of NQO1 was associated with increased FEV_1_ (**Table 1**). None of the analysed proteins showed a significant causal association with FEV_1_/FVC at FDR < 0.1. MHC class I polypeptide-related sequence B (MICB) showed a significant causal association with COPD risk (**Table 1**) with no evidence of heterogeneity (*P* > *0*.*05*) or pleiotropy (Egger MR Intercept *P* > *0*.*05*). The direction of effect was such that increased plasma MICB level was associated with reduced COPD risk (**Table 1 and Figure 3**). None of the proteins was causally associated with more than one COPD phenotype.

**Figure 3.**
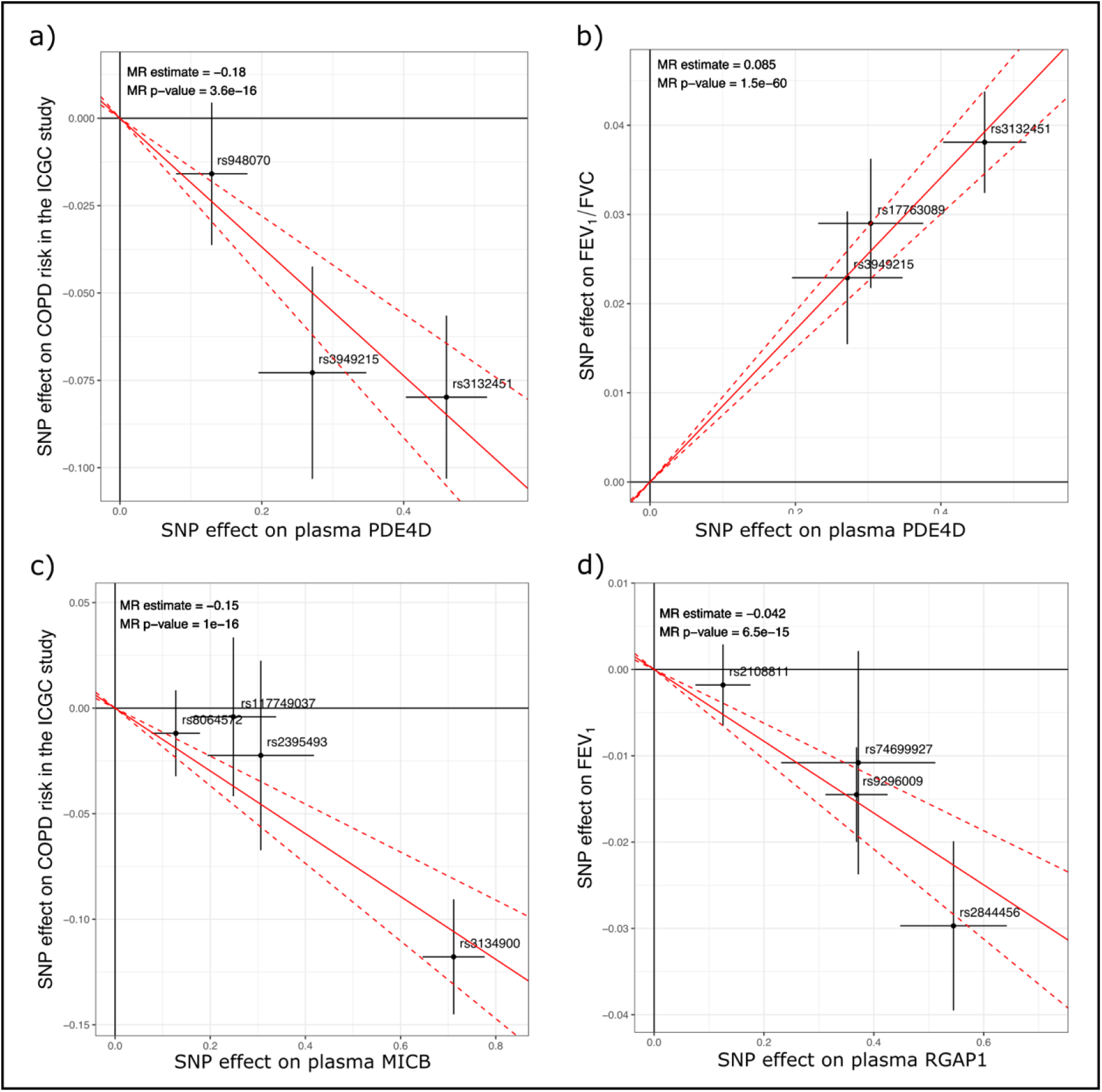
Mendelian Randomization (MR) of protein biomarkers for lung function and Chronic Obstructive Pulmonary Disease (COPD) risk. Inverse variance weighting (IVW) MR (IVW-MR) of three plasma proteins on lung function and/or COPD risk. The figure shows the IVW-MR plot for PDE4D (a and b), MICB (c) and RGAP1 (d). Dots represent the effect of the SNPs used for the IVW-MR on th plasma protein levels (horizontal axis) and lung function traits or COPD risk (vertical axis). Estimate were derived from 1) a plasma genome-wide association study (GWAS) for each protein, 2) GWAS of the International COPD Genetics Consortium (ICGC) for COPD risk and 3) a Spirometry GWAS meta-analysis (UK biobank and SpiroMeta cohorts) for the forced expiratory volume in one second (FEV_1_) and FEV_1_/Forced vital capacity (FVC). Error bars for each SNP represents the 95% confidence intervals. The slope of the red line is the instrumental variable regression estimate of the effect of protein on the lun function traits and/or COPD risk. IVW-MR *P-value* is shown at the top left corner of each MR plot.

When we compared the results of the COLOC and hypothesis-free MR, we found that three proteins (QSOX2, RGAP1 and MICB) were identified by both methods at FDR < 0.1.

### Stage 2: a step-wise discovery of plasma proteins that were causally associated with lung function and/or COPD risk

In Stage 2, we extracted the MR results for proteins showing significant colocalization (*PP*_*H4*_ >0.8) with at least one of the phenotypes (537, 607, and 250 plasma proteins that colocalized with FEV_1_, FEV_1_/FVC, and COPD risk, respectively). In this stage we considered the MR result as a complement to the COLOC results; we therefore set the significance threshold of the MR model at *P*_*MR*_ <0.05. In total, 37 unique proteins showed a causal association with at least one of the three phenotypic traits. These included 20 proteins associated with FEV_1_, 12 proteins associated with FEV_1_/FVC, and 9 proteins associated with COPD (**Table 2**). None of the significant results showed heterogeneity (*P* > *0*.*05*) or pleiotropy (Egger MR Intercept *P* > *0*.*05*).

**Table 2.**
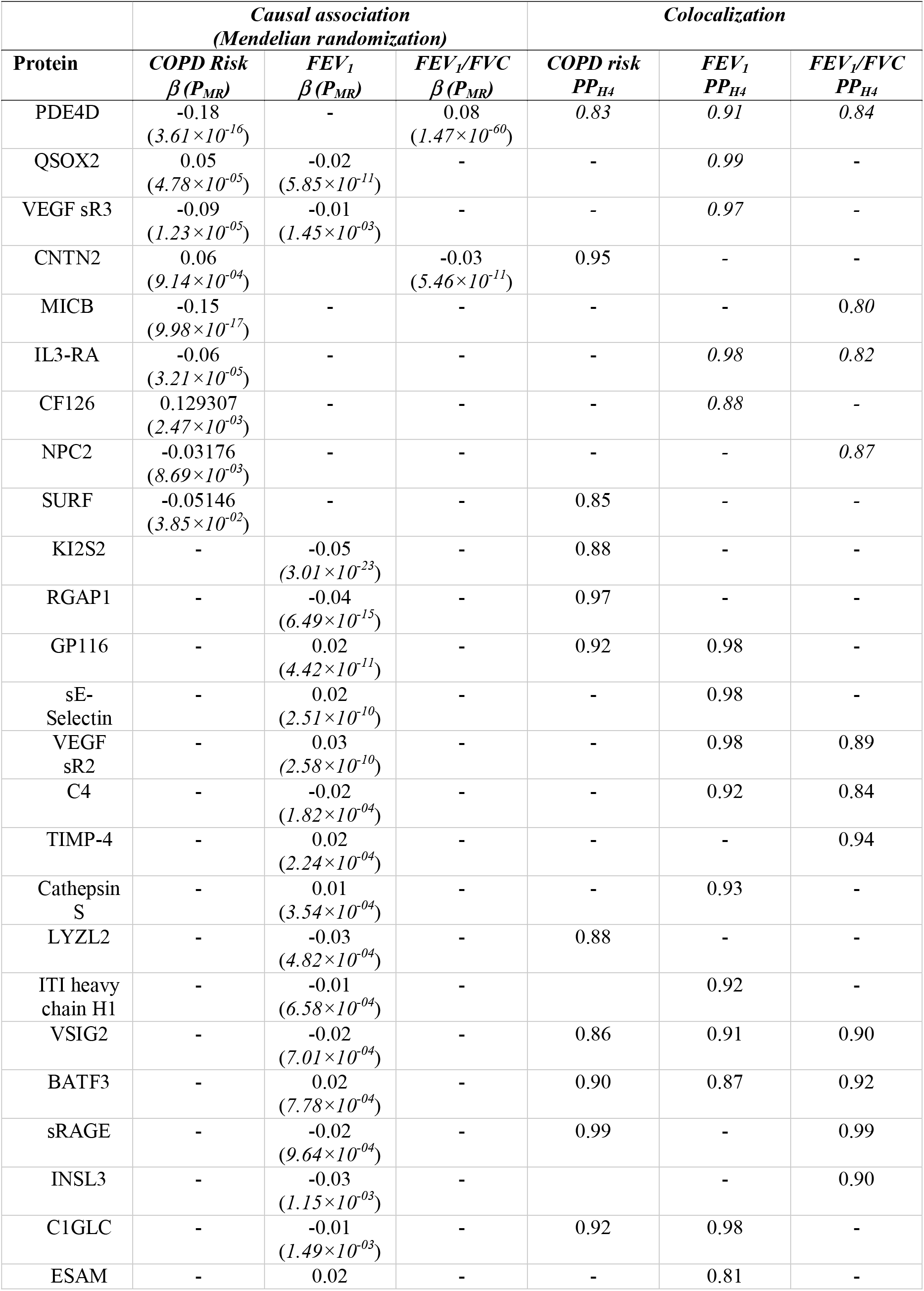
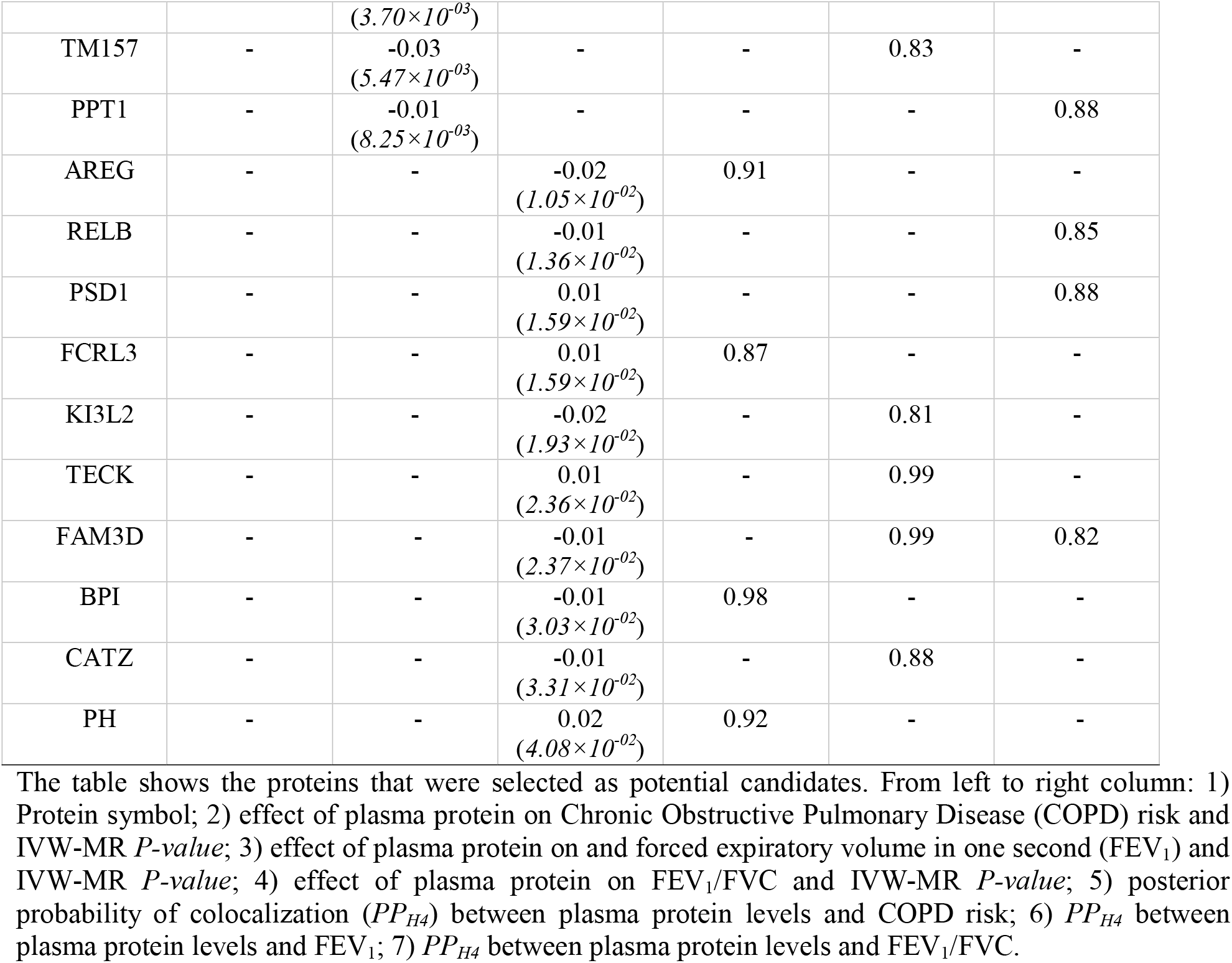
Potential plasma protein targets associated with lung function and COPD.

For FEV_1_, the most significantly associated candidate protein was RGAP1 whose plasma levels increased with decreasing FEV_1_ (**Figure 3**). For FEV1/FVC, the most significantly associated protein was cAMP-specific 3’, 5’-cyclic phosphodiesterase 4D (PDE4D), whose plasma levels increased with increasing FEV_1_/FVC (**Figure 3**). For COPD risk, the most significantly associated protein was MICB, whose plasma levels increased with decreasing COPD risk (**Figure 3**). RGAP1 and MICB, but not PDE4D, were also identified with the hypothesis-free MR approach (stage 1) with the same direction of effect for their respective phenotypic traits.

There was some overlap between the proteins associated with each of the COPD phenotypes. For example, increased plasma QSOX2 was associated with increased COPD risk and decreased FEV_1_ (**Table 2**). Increased plasma contactin-2 (CNTN2) was causally associated with decreased FEV_1_/FVC and increased COPD risk. None of the proteins tested were found to have a causal association with all three phenotypic traits.

### Highlighting potential candidate proteins

We nominated colocalized proteins having strong support for a causal association with lung function and/or COPD as candidate proteins. This list included proteins showing a significant MR estimate in Stage 2; for example, **Figure 3** shows the MR results for PDE4D, MICB and RGAP1; by nature of the step-wise analysis, these proteins were also significantly colocalized with one or more of the COPD traits. The full MR results for these proteins are provided in **Supplemental Table 2**.

The table shows the proteins that were selected as potential candidates. From left to right column: 1) Protein symbol; 2) effect of plasma protein on Chronic Obstructive Pulmonary Disease (COPD) risk and IVW-MR *P-value*; 3) effect of plasma protein on and forced expiratory volume in one second (FEV_1_) and IVW-MR *P-value*; 4) effect of plasma protein on FEV_1_/FVC and IVW-MR *P-value*; 5) posterior probability of colocalization (*PP*_*H4*_) between plasma protein levels and COPD risk; 6) *PP*_*H4*_ between plasma protein levels and FEV_1_; 7) *PP*_*H4*_ between plasma protein levels and FEV_1_/FVC.

## Discussion

Translating COPD genetics findings into actionable biomarkers and therapeutic targets requires understanding of the genes and proteins underlying the genetic associations. To our knowledge, this is the largest integrative proteomics and GWAS for lung function and COPD to date. The key findings were that: 1) the genetic loci associated with plasma levels of 1,048 and 250 unique proteins colocalized with lung function and COPD risk, respectively; 2) using an standard MR approach, we identified 4 unique plasma proteins causally associated with lung function and/or COPD risk; and 3) using an step-wise approach (COLOC coupled with MR) we identified 37 candidates proteins for lung function and/or COPD.

By integrating lung function and COPD genetics and blood proteomics, we identified 537, 607, and 250 unique proteins, whose plasma levels significantly associated with FEV_1_, FEV_1_/FVC, or COPD risk, respectively. Of these, 74 were common to all three traits. This is a substantial increase in the number of peripheral proteins associated with COPD phenotypes when compared to previous reports, which had focused on candidate genes (Obeidat et al. 2017; Milne et al. 2020). Our findings suggest that the expression levels of plasma proteins are associated with lung function and COPD risk, supporting the notion that GWAS variants (even when located within non-coding regions) have potential biological consequences that ultimately contribute to the phenotypic variation of a complex trait or disease. Our findings could be used as a starting point to support future biomarker and/or drug developmental studies. This is particular true since targets with genetic support are more likely to be successful in drug development (Nelson et al. 2015).

Of the proteins we identified in this study, there are several with known, biologically-plausible links to lung physiology and lung disease. For example, PDE4D is an isoenzyme that is part of the phosphodiesterase subfamily 4 (PDE4 A-D), a group of proteins that are involved in the pathogenesis of multiple inflammatory diseases, including but not limited to asthma and COPD. Inhibition of PDE4 increases cAMP, which appears to have a positive effect in asthma and COPD by decreasing lung inflammation (Huang and Mancini 2006) and inducing airway smooth muscle relaxation (Méhats et al. 2003). PDE4 isoforms have different physiological roles (Manning et al. 1999), and the specific role of the PDE4D isoform in lung pathology is not fully understood. Importantly, pathological contribution may be tissue specific. For example, cigarette exposure has opposite effects on PDE4D expression in alveolar compared to airway epithelial tissue (Zuo et al. 2019). In our study, PDE4D was measured in the plasma compartment, and the direction of the MR estimate suggested that increased PDE4D plasma levels were causally associated with increased lung function and reduced the risk of COPD. It is therefore plausible that a soluble form of PDE4D has different effects that the intracellular form or ones that are expressed in the local lung or immune cells. Further research is needed to understand the role of the soluble PDE4D in the pathophysiology of COPD.

Another interesting candidate we found was IL3-RA. Our results suggested that this protein shares causal loci with lung function, and that increased plasma levels of IL3-RA are associated with decreased risk of COPD. IL3-RA is one of the two subunits of the interleukin-3 (IL3) receptor, a cytokine produced mostly by T-cells and involved in several immunopathologies (Hercus et al. 2013). IL3 signalling may be important for surfactant homeostasis (Campo et al. 2012), alveolar macrophage function (Notarangelo and Pessach 2008), and activation and recruitment of eosinophils (George and Brightling 2016) (Davoine and Lacy 2014). Surfactant homeostasis appears to be a contributing factor in obstructive lung diseases, including asthma, COPD and cystic fibrosis (Devendra and Spragg 2002). Although this concept has not been fully validated by experimental studies, it is plausible that soluble IL3-RA in plasma may disrupt IL3 signalling via competitive binding inhibition. This potential function of IL3-RA in plasma may warrant further investigation especially in the context of COPD.

We found that increased sRAGE plasma protein levels were causally associated with decreased lung function. sRAGE is the soluble isoform of the receptor for advanced glycation end-products (RAGE). Under normal conditions this receptor is mainly expressed in lung tissue, particularly in type I alveolar epithelial cells, and mediates proinflammatory responses (Buckley and Ehrhardt 2010). sRAGE acts as a decoy for RAGE since it is capable of binding to RAGE ligands, therefore inhibiting RAGE (Demling et al. 2006). Dysregulation of sRAGE has been linked to COPD and lung function. Healthy individuals have been reported to have higher levels of sRAGE compared to COPD patients (Gopal et al. 2012). In contrast, our results suggest that in a general population increased sRAGE is linked to decreased lung function. This is in agreement with a study of current smokers, where a missense variant in the gene encoding for RAGE was associated with lower serum sRAGE levels and increased lung function (Miller et al. 2016). Differential effects of sRAGE may therefore depend on an individual’s genotype. To fully understand these conflicting results it is crucial to study RAGE regulation in COPD and in normal healthy individuals.

We also found that increased MICB levels were associated with decreased COPD risk. Variants within the *MICB* gene have previously been associated with lung function (Soler Artigas et al. 2011). *MICB* belongs to a major histocompatibility complex class I chain-related (MIC) gene family (Bahram 2000). MICB is a cellular stress-induced molecule that contributes to the innate and adaptive immune responses (Jamieson et al. 2002; Carapito and Bahram 2015). Soluble MICB is decreased or undetectable in bronchial washes of smokers with normal lung function and COPD patients compared to those of never-smokers (Roos-Engstrand et al. 2010). This is in agreement with the direction of the MICB effect suggested by our study, and may therefore be an interesting plasma protein candidate for further studies.

Our study has a number of limitations. First, the cohorts used for our analysis only included white-European individuals; therefore the conclusions based on our results may not be generalisable to populations of different ancestry. Second, our study was limited to the blood plasma proteome. Since COPD is a systemic disease, studying the proteome of other tissues and cell types may further elucidate the mechanisms underlying the GWAS associations. Third, the 2,995 proteins found in the INTERVAL Study data set represent ∼15% of the whole human proteome. It is probable that proteins not measured on this platform may contribute to the phenotypic variability of the traits; these proteins remain undiscovered. Furthermore, it is likely that other mechanisms that we did not explore (e.g. epigenetics and gene expression) could contribute to COPD risk. Fourth, our integrative-omics approach only explored cis regions, therefore future research should also evaluate the trans or distal regions associated with the traits. Lastly, although our approach allowed us to prioritize a manageable set of proteins, further efforts are necessary to validate the role of these peripheral proteins and their relationship to lung function and COPD.

In summary, our integrative-omics approach revealed several novel plasma proteins that were significantly linked with COPD risk and/or lung function. Using a MR framework, we provide evidence suggesting that the plasma levels of multiple proteins have causal effects on these phenotypic traits. These proteins represent promising candidates for future development of biomarkers and/or therapeutic targets in COPD and other lung pathologies associated with reduced lung function.

## Materials and methods

### Datasets analysed for the current study

#### INTERVAL study

We analysed plasma protein quantitative trait loci (pQTL) obtained from the INTERVAL study. The INTERVAL study was a randomized trial of blood donation intervals that comprises around 45,000 participants that were recruited between June 11, 2012, and June 15, 2014 (Di Angelantonio et al. 2017; Sun et al. 2018). The full details on the criteria for participants’ recruitment, informed consent, description of the cohort, sample collection, INTERVAL study design, and objectives have been previously published (Di Angelantonio et al. 2017; Sun et al. 2018). Briefly, participants from the INTERVAL study were aged 18 years and older, in general good health (based on blood donation criteria), and were recruited at 25 static donor centres of NHS Blood and Transplant (NHSBT). Blood collection was performed using standard venepuncture. Participants were genotyped for about 830,000 genetic variants using the Affymetrix Axiom UK Biobank genotyping array and imputed to the 1000 Genome phase 3 UK10K reference panel. Genetic variants with imputation score (r^2^) >0.7, Hardy-Weinberg Equilibrium (HWE) *P*>*5*×*10*^-06^ and minor allele count of > 8 were retained (10,572,788 genetic variants). A full description of the genotyping protocol and quality control has been previously described (Astle et al. 2016).

After filtering out participants who failed to pass the genetic quality controls (HWE, minor allele count, r^2^), a randomly-selected subset of 3,301 participants was used for the plasma pQTL analyses of 2,995 plasma proteins levels. The protein levels were log-transformed and adjusted for age, sex, waiting period between blood collection and processing and the first three genetic principal components. The protein residuals then were extracted and rank-inverse normalized. Later these residuals were used for genome-wide associations study (GWAS). The genetic associations were tested with linear regression using an additive genetic model and the results from each cohort were combined using a fixed-effect inverse-variance meta-analysis. Significant associations were defined at *P*<*1*.*5*×*10*^*− 11*^. We used the totality of the pQTL results (summary statistics) for the subsequent analyses described later in this section.

#### International COPD Genetics Consortium (ICGC) study

We determined the relationship between plasma proteins and COPD risk using the ICGC dataset. The ICGC study is one of the largest COPD GWASs that has been conducted to date. Briefly, this cohort included over 200,000 participants (35,735 cases and 222,076 controls) from individual COPD GWASs (Sakornsakolpat et al. 2019). Each individual study obtained informed consent from individual participants, and approval from the local human research ethics/regulatory bodies (Cho et al. 2014; Hobbs et al. 2017; Sakornsakolpat et al. 2019). COPD cases were defined based on pre-bronchodilator spirometry, with FEV_1_ < 80% predicted and FEV_1_ to FVC ratio of < 0.70. Controls were defined as FEV_1_ > 80% predicted and FEV_1_/FVC > 0.70. Each COPD GWAS was evaluated using logistic regression, which adjusted for age, sex, pack-year of smoking, ever smoking status, current smoking status, and genetic ancestry (principal component, as required for each study). The GWAS results were combined using a fixed-effects meta-analysis. A full description of the cohort and study methods have been previously published (Cho et al. 2014; Hobbs et al. 2017; Sakornsakolpat et al. 2019).

#### UK biobank and SpiroMeta lung function meta-analysis

We used genome-wide associations with lung function (FEV_1_ and FEV_1_/FVC) from a previously-published meta-analysis of the UK Biobank and SpiroMeta datasets (Shrine et al. 2019). This meta-analysis included 321,047 and 79,055 white European participants from the UK Biobank project (Bycroft et al. 2018) and the SpiroMeta consortium, respectively.

In the UK Biobank, phenotypic information was collected in 22 recruitment centres across the United Kingdom (Bycroft et al. 2018). Human research ethics approval for the UK Biobank project was granted by the North West Multi-centre Research Ethics Committee (MREC). Participants were genotyped with the Affymetrix Axiom UK BiLEVE and UK biobank array (Wain et al. 2015) and imputed to the Haplotype Reference Consortium panel; genotypes were retained if the minor allele count was ≥ 3 and imputation r^2^>0.5 (Bycroft et al. 2018). Association testing was conducted between genome-wide-significant genotypes and lung function traits (FEV_1_ and FEV_1_/FVC) using a linear mixed model (LMM), assuming additive genetic effects (Loh et al. 2015). The LMM was adjusted for confounders (age, age^2^, sex, height, smoking status, and genotyping array) (Shrine et al. 2019).

The SpiroMeta consortium includes participants from 22 individual studies, which have been previously described (Shrine et al. 2019). Approval was granted for each individual study from their respective local human research ethics committees. Participants were genotyped according to the protocol described in each of the studies; 13 studies were imputed to the 1000 Genomes Project Phase 1 panel (1000 Genomes Project Consortium et al. 2010) and nine to the Haplotype Reference Consortium panel (McCarthy et al. 2016). Genotypes with imputation r^2^<0.30 were excluded from further analysis (Shrine et al. 2019). For each individual study, genetic associations with FEV_1_ and FEV_1_/FVC were determined using a linear regression model adjusted for age, age^2^, sex and height. Genetic principal components were included as covariates in studies of unrelated participants, while LMM were used in studies of related participants to account for kinship and population structure. The results across all studies in the SpiroMeta consortium were combined using an inverse-variance weighted meta-analysis. Shrine and colleagues (Shrine et al. 2019) then combined the UK Biobank and SpiroMeta GWAS results using an inverse-variance-weighted fixed-effects meta-analysis. In total, 19,819,130 genetic variants (imputed or genotyped) in both cohorts (UK biobank and SpiroMeta consortium) were used for the meta-analysis. The LD regression score intercept was close to 1(Shrine et al. 2019), therefore no genomic control was applied.

### Integrative -omics methods

#### Bayesian colocalization (COLOC)

We used COLOC to determine whether the associations between lung function traits (FEV_1_, FEV_1_/FVC, and COPD risk) and plasma protein levels were consistent with a causal variant (colocalization). We performed the analysis using the coloc package (Giambartolomei et al. 2014) implemented in R (R Core Team 2018). We evaluated the summary statistics for 3,248 plasma pQTLs (Sun et al. 2018) from the INTERVAL study. We included all SNPs associated with lung function (FEV_1_ or FEV_1_/FVC) in the UK Biobank/SpiroMeta GWAS meta-analysis (19,819,130) (Shrine et al. 2019) and all SNPs associated with COPD risk in the ICGC dataset (6,224,355). In order to maximise the number of variants tested per genomic locus, which overlapped across these studies, we did not set an *a priori* p value threshold for inclusion.

The COLOC method used for our research calculates the posterior probability (*PP*) of colocalization of lung function (or COPD risk) and plasma protein-associated variants within a defined genomic region. In summary, a large *PP* is supportive evidence of a single shared causal variant for the two traits. For this test we applied the following parameters: 1) a prior probability of a SNP being associated with the lung function trait or plasma protein level to 1×10^−04^; and 2) a prior probability of a SNP being associated with the lung function trait and plasma protein level to 1×10^−06^. All SNPs across the genome were assumed to have equal prior probabilities (as set before). In addition, we defined loci to be tested as genomic regions ± 0.5 Mb windows surrounding the top lung function-associated (or COPD risk-associated) variants. As a result, we applied COLOC to 988,183 loci for FEV_1_, 1,109,654 for FEV_1_/FVC and 282,338 for COPD risk. We placed the significance threshold of the COLOC analysis to be *PP*_*H4*_>0.80.

#### Mendelian Randomization

We used a MR approach to identify causal relationships between plasma proteins (“exposure”) and the three traits (FEV_1_, FEV_1_/FVC, and COPD risk; “traits”). We first identified pQTLs for each of the selected proteins in the INTERNAL study by extracting the effect size (Beta) and standard error (SE) of each variant that at least reached the arbitrary threshold of *P***<** *5*×*10*^*-06*^ and excluded the variants that were found within a 2Mb window(± 1Mb) to avoid linkage. Next, we examined the complex trait associations in the UK Biobank/SpiroMeta meta-analysis (FEV_1_, FEV_1_/FVC) and ICGC (COPD risk) for these variants and, if present, extracted the Beta value and SE for each. We then used these genetic variants as instrumental variables (IVs) in a MR analysis. The fundamental assumptions of MR analysis are: 1) that the IVs are associated with the exposure; 2) that the selected IVs only affect an outcome *via* the exposure; and 3) that the IVs are independent of confounders. Using two MR methods, inverse variance weighting (IVW) MR (IVW-MR) and Egger-MR we aimed to identify causal risk factors. For each exposure-outcome pairing, we used an inverse variance weighted linear regression model (IVW-MR) to relate the per-allele SNP association with the exposure to its association with the trait. IVW-MR assumes no directional pleiotropy (i.e. genetic variant associated with multiple unrelated phenotypes) by constraining the intercept to zero, and accounts for linkage disequilibrium (LD) between genetic variants, heterogeneity was assessed based on the Cochran’s Q test (*P*<*0*.*05*) that is part of the IVW-MR outcome (Burgess et al. 2016). We also performed Egger-MR, which accounts for directional pleiotropy by un-constraining the intercept (Bowden et al. 2015).

Based on this workflow, we identified a protein as having a causal association with the complex trait if the IVW-MR estimate was significant *FDR* < *0*.*1* and the Egger-MR intercept was not different from zero (*P* > *0*.*05*). In addition, using an *a priori* hypothesis that the colocalized proteins were causally related to lung function traits and/or COPD, we determined their significant causal associations based on the IVW-MR threshold of *P*_*MR*_ <*0*.*05* and Egger-MR intercept *P* > *0*.*05*.

#### Potential Biomarkers

Using the workflow described in **Figure 1**, we aimed to develop a list of potential candidate proteins that warrant further investigation for their role in lung function and/or COPD pathogenesis. We selected as top plasma proteins those that showed both significant colocalization at *PP*_*H4*_ > 0.80 and *P*_*MR*_ < 0.05 with lung function and/or COPD risk.

## Data Availability

All data used for these analyses are publicly available.

https://www.ebi.ac.uk/gwas/publications/30804560

http://www.phpc.cam.ac.uk/ceu/proteins/

https://www.ebi.ac.uk/gwas/publications/30804561

## Article information

### Data availability

All data used for these analyses are publicly available at: http://www.phpc.cam.ac.uk/ceu/proteins/; https://www.ebi.ac.uk/gwas/publications/30804560; and https://www.ebi.ac.uk/gwas/publications/30804561.

## Acknowledgement

Compute Canada computer cluster was used to conduct the data analyses.

## References

1000 Genomes Project Consortium, Abecasis GR, Altshuler D, et al (2010) A map of human genome variation from population-scale sequencing. Nature 467:1061–1073. https://doi.org/10.1038/nature09534

Astle WJ, Elding H, Jiang T, et al (2016) The Allelic Landscape of Human Blood Cell Trait Variation and Links to Common Complex Disease. Cell 167:1415-1429.e19. https://doi.org/10.1016/j.cell.2016.10.042

Bahram S (2000) MIC genes: from genetics to biology. Adv Immunol 76:1–60. https://doi.org/10.1016/s0065-2776(01)76018-x

Bowden J, Davey Smith G, Burgess S (2015) Mendelian randomization with invalid instruments: effect estimation and bias detection through Egger regression. Int J Epidemiol 44:512– 525. https://doi.org/10.1093/ije/dyv080

Buckley ST, Ehrhardt C (2010) The receptor for advanced glycation end products (RAGE) and the lung. J Biomed Biotechnol 2010:917108. https://doi.org/10.1155/2010/917108

Burgess S, Dudbridge F, Thompson SG (2016) Combining information on multiple instrumental variables in Mendelian randomization: comparison of allele score and summarized data methods. Stat Med 35:1880–1906. https://doi.org/10.1002/sim.6835

Bycroft C, Freeman C, Petkova D, et al (2018) The UK Biobank resource with deep phenotyping and genomic data. Nature 562:203–209. https://doi.org/10.1038/s41586-018-0579-z

Campo I, Kadija Z, Mariani F, et al (2012) Pulmonary alveolar proteinosis: diagnostic and therapeutic challenges. Multidisciplinary Respiratory Medicine 7:4. https://doi.org/10.1186/2049-6958-7-4

Carapito R, Bahram S (2015) Genetics, genomics, and evolutionary biology of NKG2D ligands. Immunol Rev 267:88–116. https://doi.org/10.1111/imr.12328

Cho MH, McDonald M-LN, Zhou X, et al (2014) Risk loci for chronic obstructive pulmonary disease: a genome-wide association study and meta-analysis. Lancet Respir Med 2:214– 225. https://doi.org/10.1016/S2213-2600(14)70002-5

Davoine F, Lacy P (2014) Eosinophil cytokines, chemokines, and growth factors: emerging roles in immunity. Front Immunol 5:570. https://doi.org/10.3389/fimmu.2014.00570

Demling N, Ehrhardt C, Kasper M, et al (2006) Promotion of cell adherence and spreading: a novel function of RAGE, the highly selective differentiation marker of human alveolar epithelial type I cells. Cell Tissue Res 323:475–488. https://doi.org/10.1007/s00441-005-0069-0

Devendra G, Spragg RG (2002) Lung surfactant in subacute pulmonary disease. Respiratory Research 3:11. https://doi.org/10.1186/rr168

Di Angelantonio E, Thompson SG, Kaptoge S, et al (2017) Efficiency and safety of varying the frequency of whole blood donation (INTERVAL): a randomised trial of 45L000 donors. Lancet 390:2360–2371. https://doi.org/10.1016/S0140-6736(17)31928-1

George L, Brightling CE (2016) Eosinophilic airway inflammation: role in asthma and chronic obstructive pulmonary disease. Ther Adv Chronic Dis 7:34–51. https://doi.org/10.1177/2040622315609251

Giambartolomei C, Vukcevic D, Schadt EE, et al (2014) Bayesian test for colocalisation between pairs of genetic association studies using summary statistics. PLoS Genet 10:e1004383. https://doi.org/10.1371/journal.pgen.1004383

Gopal P, Rutten EPA, Dentener MA, et al (2012) Decreased plasma sRAGE levels in COPD: influence of oxygen therapy. Eur J Clin Invest 42:807–814. https://doi.org/10.1111/j.1365-2362.2012.02646.x

Gusev A, Ko A, Shi H, et al (2016) Integrative approaches for large-scale transcriptome-wide association studies. Nature Genetics 48:245–252. https://doi.org/10.1038/ng.3506

Hercus TR, Dhagat U, Kan WLT, et al (2013) Signalling by the βc family of cytokines. Cytokine Growth Factor Rev 24:189–201. https://doi.org/10.1016/j.cytogfr.2013.03.002

Hobbs BD, de Jong K, Lamontagne M, et al (2017) Genetic loci associated with chronic obstructive pulmonary disease overlap with loci for lung function and pulmonary fibrosis. Nat Genet 49:426–432. https://doi.org/10.1038/ng.3752

Huang Z, Mancini JA (2006) Phosphodiesterase 4 inhibitors for the treatment of asthma and COPD. Curr Med Chem 13:3253–3262. https://doi.org/10.2174/092986706778773040

Jamieson AM, Diefenbach A, McMahon CW, et al (2002) The role of the NKG2D immunoreceptor in immune cell activation and natural killing. Immunity 17:19–29. https://doi.org/10.1016/s1074-7613(02)00333-3

Lamontagne M, Bérubé J-C, Obeidat M, et al (2018) Leveraging lung tissue transcriptome to uncover candidate causal genes in COPD genetic associations. Hum Mol Genet 27:1819– 1829. https://doi.org/10.1093/hmg/ddy091

Loh P-R, Tucker G, Bulik-Sullivan BK, et al (2015) Efficient Bayesian mixed-model analysis increases association power in large cohorts. Nat Genet 47:284–290. https://doi.org/10.1038/ng.3190

Manning CD, Burman M, Christensen SB, et al (1999) Suppression of human inflammatory cell function by subtype-selective PDE4 inhibitors correlates with inhibition of PDE4A and PDE4B. Br J Pharmacol 128:1393–1398. https://doi.org/10.1038/sj.bjp.0702911

McCarthy S, Das S, Kretzschmar W, et al (2016) A reference panel of 64,976 haplotypes for genotype imputation. Nat Genet 48:1279–1283. https://doi.org/10.1038/ng.3643

Méhats C, Jin S-LC, Wahlstrom J, et al (2003) PDE4D plays a critical role in the control of airway smooth muscle contraction. FASEB J 17:1831–1841. https://doi.org/10.1096/fj.03-0274com

Miller S, Henry AP, Hodge E, et al (2016) The Ser82 RAGE Variant Affects Lung Function and Serum RAGE in Smokers and sRAGE Production In Vitro. PLoS One 11:. https://doi.org/10.1371/journal.pone.0164041

Milne S, Li X, Cordero AIH, et al (2020) Protective effect of club cell secretory protein (CC-16) on COPD risk and progression: a Mendelian randomisation study. Thorax 75:934–943. https://doi.org/10.1136/thoraxjnl-2019-214487

Nelson MR, Tipney H, Painter JL, et al (2015) The support of human genetic evidence for approved drug indications. Nature Genetics 47:856–860. https://doi.org/10.1038/ng.3314

Notarangelo LD, Pessach I (2008) Out of breath: GM-CSFRα mutations disrupt surfactant homeostasis. Journal of Experimental Medicine 205:2693–2697. https://doi.org/10.1084/jem.20082378

Obeidat M, Hao K, Bossé Y, et al (2015) Molecular mechanisms underlying variations in lung function: a systems genetics analysis. Lancet Respir Med 3:782–795. https://doi.org/10.1016/S2213-2600(15)00380-X

Obeidat M, Li X, Burgess S, et al (2017) Surfactant protein D is a causal risk factor for COPD: results of Mendelian randomisation. European Respiratory Journal 50:. https://doi.org/10.1183/13993003.00657-2017

R Core Team (2018) R: A language and environment for statistical computing. R Foundation for Statistical Computing. https://www.r-project.org/. Accessed 5 Sep 2019

Roos-Engstrand E, Pourazar J, Behndig AF, et al (2010) Cytotoxic T cells expressing the costimulatory receptor NKG2 D are increased in cigarette smoking and COPD. Respiratory Research 11:128. https://doi.org/10.1186/1465-9921-11-128

Roth GA, Abate D, Abate KH, et al (2018) Global, regional, and national age-sex-specific mortality for 282 causes of death in 195 countries and territories, 1980–2017: a systematic analysis for the Global Burden of Disease Study 2017. The Lancet 392:1736– 1788. https://doi.org/10.1016/S0140-6736(18)32203-7

Sakornsakolpat P, Prokopenko D, Lamontagne M, et al (2019) Genetic landscape of chronic obstructive pulmonary disease identifies heterogeneous cell-type and phenotype associations. Nat Genet 51:494–505. https://doi.org/10.1038/s41588-018-0342-2

Shrine N, Guyatt AL, Erzurumluoglu AM, et al (2019) New genetic signals for lung function highlight pathways and chronic obstructive pulmonary disease associations across multiple ancestries. Nat Genet 51:481–493. https://doi.org/10.1038/s41588-018-0321-7

Smith GD, Ebrahim S (2003) “Mendelian randomization”: can genetic epidemiology contribute to understanding environmental determinants of disease? Int J Epidemiol 32:1–22. https://doi.org/10.1093/ije/dyg070

Soler Artigas M, Loth DW, Wain LV, et al (2011) Genome-wide association and large-scale follow up identifies 16 new loci influencing lung function. Nat Genet 43:1082–1090. https://doi.org/10.1038/ng.941

Sun BB, Maranville JC, Peters JE, et al (2018) Genomic atlas of the human plasma proteome. Nature 558:73–79. https://doi.org/10.1038/s41586-018-0175-2

Voight BF, Peloso GM, Orho-Melander M, et al (2012) Plasma HDL cholesterol and risk of myocardial infarction: a mendelian randomisation study. Lancet 380:572–580. https://doi.org/10.1016/S0140-6736(12)60312-2

Wain LV, Shrine N, Miller S, et al (2015) Novel insights into the genetics of smoking behaviour, lung function, and chronic obstructive pulmonary disease (UK BiLEVE): a genetic association study in UK Biobank. Lancet Respir Med 3:769–781. https://doi.org/10.1016/S2213-2600(15)00283-0

Zuo H, Faiz A, van den Berge M, et al (2019) Cigarette smoke exposure alters phosphodiesterases in human structural lung cells. American Journal of Physiology-Lung Cellular and Molecular Physiology 318:L59–L64. https://doi.org/10.1152/ajplung.00319.2019

